# Early detection of hip dysplasia by nurse-led ultrasound screening during home visits: A preliminary prospective cohort study

**DOI:** 10.64898/2026.04.07.26350299

**Authors:** Kyoko Yoshioka-Maeda, Hiroshige Matsumoto, Chikako Honda, Takeshi Kinjo, Kiyoshi Aoki, Keita Okada, Kenta Fujiwara

**Author notes:** Correspondence concerning this article should be addressed to: Kyoko Yoshioka-Maeda, Department of Community Health Nursing, Division of Health Sciences and Nursing, Graduate School of Medicine, The University of Tokyo, 7-3-1 Hongo, Bunkyo-ku, Tokyo 113-0033, Japan. **E-mail address for all authors**. **Statements Data availability statement**: Research data are not publicly shared because of privacy restrictions. **Conflict of interest disclosure:** The authors declare no conflict of interest. The pediatric orthopedic surgeons (TK, KA, and KF) are board members and instructors of the Japanese Society of Orthopedic Ultrasonics. **Ethics approval statement:** The primary researcher’s Institutional Review Board (IRB) approved the study protocol (Research Ethics Committee of the Faculty of Medicine of the University of Tokyo, no. 2022310NI, 22 February 2023; 2022310NI-(1), 14 June 2023; no. 2022310NI-(2), 12 June 2024; 20223101NI; 28 July 2023; 2023101NI-(1),12 June 2024). The ultrasound hip screening was implemented as part of the local government’s newborn and infant HV program. The study was explained to each caregiver using documents and a website that the data might be used for research after personal information had been removed. Caregivers were provided with detailed information about the study through written documents and a dedicated website, which explained that anonymized data from the screenings might be used for research purposes. A six-week opt-out period followed each visit, and data were included in the study only after this period had elapsed. No caregivers opted out during the designated period, and all data were included in the analysis. **Patient consent statement:** No caregivers opted out during the designated period. **Permission to reproduce material from other sources:** Not applicable. **Clinical trial registration:** The protocol of this study was registered in the University Hospital Medical Information Network Clinical Trial Registry before starting the study (no. UMIN000051929, 16 August 2023). **Acknowledgments:** The authors thank all the public health nurses, relevant staff, newborns, infants, and caregivers who supported this study. We thank Dr. Keiichi Nakagawa (Department of Bioengineering, Department of Precision Engineering, School of Engineering, The University of Tokyo); Dr. Naoki Tomii, Dr. Masayoshi Tsuzuki, and Mr. Ryosuke Oki (Biomedical Precision Engineering Laboratory, Department of Precision Engineering, Faculty of Engineering, The University of Tokyo); Professor Dr. Gojiro Nakagami, Dr. Mari Abe-Doi, and Professor Emeritus Dr. Hiromi Sanada (Department of Gerontological Nursing/Wound Care Management, Division of Health Sciences and Nursing, Graduate School of Medicine, The University of Tokyo; President of Ishikawa Prefectural Nursing University); Professor Dr. Megumi Haruna (Department of Midwifery and Women’s Health, Division of Health Sciences and Nursing, Graduate School of Medicine, The University of Tokyo); Dr. Toshiaki Takahashi (School of Medicine Nursing Course Adult Nursing, Yokohama City University), Dr. Kazuyuki Komichi (RINGNE); and The Okinawa Association of Child Health for their invaluable contributions to our project. **Author contributions:** All authors met the criteria of the International Committee for Medical Journal Editors through direct contributions to the design and conduct of this study. All authors had significantly revised the draft and confirmed the final version of the manuscript. Conceptualization: K.Y.-M., H.M., C.H., T.K., K.A., and K.F.; methodology: K.Y.-M., C.H., H.M. T.K., K.A., O.K., and K.F.; software: H.M.; validation: K.Y.-M., H.M., C.H., T.K., K.A., and K.F.; resources: K.Y.-M., H.M., C.H., T.K., K.A., O.K., and K.F.; data curation: K.Y.-M., H.M., and C.H.; writing—original draft preparation: K.Y.-M. and H.M.; writing—review and editing: K.Y.-M., H.M., C.H., T.K., K.F., O.K., and K.A.; visualization: H.M. and C.H.; supervision: K.Y.-M., T.K., K.F., K.O., and K.A.; project administration, K.Y.-M.; and funding acquisition: K.Y.-M. All the authors have read and agreed to the published version of the manuscript.

## Abstract

**Objective:** To evaluate the feasibility of nurse-led ultrasound hip screening for newborns and infants during home visits, focusing on whether trained public health nurses (PHNs) can obtain interpretable images for orthopedic pediatric surgeons’ diagnosis, imaging error patterns, immediate operational challenges, and follow-up results of infants with suspected developmental dysplasia of the hip (DDH).

Design

Pilot prospective cohort study.

**Sample:** Forty-two infants were screened, PHNs conducted ultrasound hip screenings during home visits.

**Measurements:** Diagnostically interpretable images, as determined by two pediatric orthopedic surgeons.

**Results:** Diagnostically interpretable images of 75/84 (89.3%) hips were obtained. Surgeons identified three error patterns: incomplete visualization of the ilium (n = 2), joint capsule (n = 1), or bony roof (n = 2). Infant crying was an operational challenge (n = 1). Thirty-three (78.6%) hips were normal, four (9.5%) had abnormal findings requiring abduction exercises, three (7.1%) were referred to a hospital, and two (4.8%) failed imaging. One hip was diagnosed with subluxation, which went undetected by physical or risk screening.

**Conclusion:** Nurse-led ultrasound hip screening for newborns and infants during home visits is feasible and may aid in early DDH detection. Further studies should assess diagnostic accuracy, cost-effectiveness, and long-term outcomes.

## 1. Background

Early detection of developmental dysplasia of the hip (DDH) is essential to allow timely intervention, prevent the development of ambulatory impairments that significantly affect quality of life, ensure lifelong mobility in children, and avoid the need for hospitalization and surgical interventions (Agostiniani et al., 2020; Broadhurst, 2019; Lucchesi et al., 2021; Sato et al., 2024; Shorter et al., 2013). To detect DDH, physicians typically conduct physical examinations to identify click signs, Allis signs, thigh-crease asymmetry, and limited abduction (Harper et al., 2020), while considering risk factors such as race, sex, family history, birth order, breech presentation, oligohydramnios, birth season, cultural norms, and swaddling practices (Loder & Skopelja, 2011; Manoukian & Rehm, 2019; Shorter et al., 2013; Vaidya et al., 2021). Delayed DDH diagnosis can result in gait disabilities in children and poses a significant public-health concern (Davies et al., 2020; Hattori et al., 2017). Therefore, improving screening accuracy is critical for early DDH detection in community settings.

Early DDH detection, which is influenced by both genetic and environmental factors (Ishida, 1977). The risk of hip dislocation increases when the lower limbs are extended for prolonged periods; maintaining the feet in a natural M shape is a crucial preventive measure (Vaidya et al., 2021). Traditionally, hip dislocation is detected through physical assessments—e.g., the Ortolani and Barlow tests—and clinical observations—e.g., asymmetry in leg length and thigh creases (Harper et al., 2020). Although straightforward for physicians and nurses, the reliability of these assessments’ findings are highly dependent on experience (Harper et al., 2020; Smart et al., 2024a; Smart et al., 2024b); additionally, the detection of bilateral dislocations and early acetabular dysplasia can be challenging.

Even though the criteria for late diagnosis vary across countries, delayed DDH detection has consistently been reported to lead to walking abnormalities in children (Broadhurst, 2019; Hattori et al., 2017). Thus, improving hip-screening accuracy is critical for ensuring lifelong mobility.

Ultrasonography, which allows the visualization of newborns’ and infants’ cartilage without radiation exposure, is one of the most effective tools for detecting DDH (Agostiniani et al., 2020; Duarte et al., 2022). Despite this procedure’s costs, there is a strong international consensus on the need for universal ultrasound hip screening (Agostiniani et al., 2020), as selective screening cannot provide early diagnoses in the absence of DDH risk factors (Broadhurst, 2019). For over four decades, ultrasound hip screenings have been conducted in hospitals or clinical settings by pediatric orthopedic surgeons (Graf, 1980). Ultrasound professionals vary by country and healthcare system and may include orthopedic specialists, pediatricians, and radiologists (Krysta et al., 2024). Recently, nurse-led ultrasound screening has been reported as a feasible approach in hospital settings (Ulziibat et al., 2024). However, evidence for its feasibility in community settings remains limited (Yoshioka-Maeda et al., 2024).

Home visits (HVs) by nurses are widely recognized as the best practice for promoting maternal and child health (Olds et al., 2019; World Health Organization, 2015). However, unlike controlled clinical environments, home settings are diverse and complex (Zarrabi et al., 2022), and hip-screening accuracy can vary depending on nurses’ skill and experience (Smart et al., 2024). Therefore, a standardized ultrasound hip-screening procedure may improve early DDH detection in community settings.

Nurses already perform physical assessments of newborns and infants during HVs; training nurses in ultrasonography, in addition to manual assessments for DDH detection, can be an efficient, effective, and practical approach. In Japan, nurse-led universal ultrasound hip screening of newborns and infants during HVs has recently been implemented (Yoshioka-Maeda et al., 2024). However, whether this approach allows for effective and early DDH detection remains to be determined.

This formative pilot study aimed to evaluate the feasibility of nurse-led ultrasound hip screening of newborns and infants during HVs, focusing on whether trained public health nurses (PHNs) can obtain interpretable images that enable diagnoses by orthopedic pediatric surgeons, and what the imaging error patterns, immediate operational challenges, and the follow-up results of infants suspected of DDH for orthopedic pediatric surgeons’ review are.

## 2. Methods

### 2.1. Design

This was a prospective cohort study.

### 2.2. Sample

This study was conducted between March and August 2024 in a municipality with an annual birth rate of approximately 110 individuals. Six PHNs conducted HVs for households with infants and newborns. The nurses completed approximately 105 min of e-learning on the Graf method of hip ultrasonography (Graf, 1980; Graf et al., 2013), followed by a two-day practical training course. Educational materials and practical training were supervised by pediatric orthopedic surgeons (TK, KA, and KF) who are board members and instructors of the Japanese Society of Orthopedic Ultrasonics. Ultrasound hip screening was integrated into the existing newborn and infant HV program. As all newborns were eligible for the program, no sample-size calculation was required. During HVs, caregivers were asked if they consented to ultrasound hip screening.

### 2.3. Inclusion and Exclusion Criteria

The inclusion criteria were: 1) participation in the municipality’s newborn HV program and 2) caregiver’s consent for ultrasound hip screening. Children whose caregivers declined ultrasound hip screening were excluded.

### 2.4. Intervention

After obtaining caregivers’ informed consent, trained PHNs administered ultrasound hip screenings along with to standard physical assessments. To ensure safety in varied home environments, ultrasound hip screenings were performed on the floor (Yoshioka-Maeda et al., 2023). All caregivers were also provided with preventive health guidance on DDH in the form of pamphlets from the Japanese Society of Pediatric Orthopedic Surgeons (Japanese Pediatric Orthopaedic Association).

### 2.5. Validity and Feasibility of the Intervention

All PHNs passed the Objective Structured Clinical Examination. The Graf method uses standardized procedures to ensure accuracy (Chavoshi et al., 2021; Duarte et al., 2022; Graf, 1980; Graf et al., 2013); if the position of the child or the handling of the probe was incorrect, a standard image could not be captured (Graf et al., 2013). To ensure correct positioning, the PHNs and caregivers placed the child between two rolled-up bath towels (Figure 1). The PHN identified the greater trochanter with their index and middle fingers and placed the ultrasound probe vertically over it. Recording began as soon as screening started and stopped once the standard image—including the three landmarks of the lower limb of the ilium, a clear and straight echo of the mid-bony roof, and the joint capsule—was captured (Graf, 1980). The right hip was examined first, followed by the left, using the same procedure for both (Graf, 1980).

**Figure 1.**
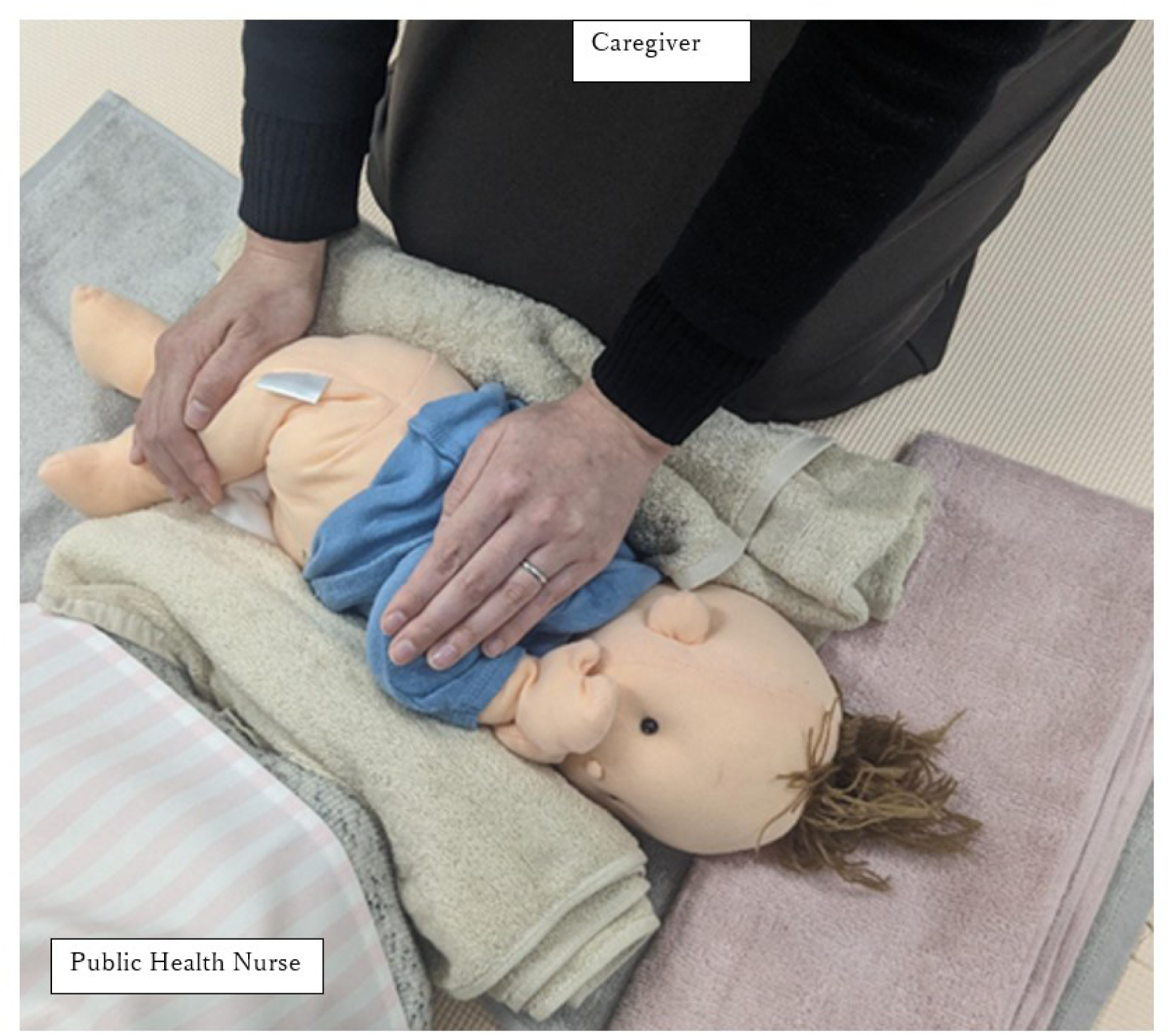
The setting of ultrasound hip screening at a home visit

Ultrasound images’ quality was ensured through a two-step process involving a real-time technical-quality check and post-hoc diagnostic-quality validation. First, during each HV, PHNs were required to obtain a standard Graf image within three minutes, based on their training (Yoshioka-Maeda et al., 2025), as the recording function of the ultrasound device allows a maximum of three minutes of continuous video capture. If the standard landmarks—the lower limb of the ilium, mid-bony roof, and joint capsule—were not clearly visualized within this timeframe, the image was immediately re-recorded. Furthermore, the PHNs recorded field notes explaining any difficulties encountered during imaging. These qualitative data were also analyzed with the surgeons’ comments to explore the technical and operational feasibility of this approach. Ultrasound images and anonymized data were subsequently uploaded to Google Cloud Storage (Google LLC, Mountain View, CA, USA) for interpretation by pediatric orthopedic surgeons.

Second, after data were uploaded, a pediatric orthopedic surgeon (TK) independently reviewed all images to verify whether each was diagnostically interpretable and met the standard plane criteria defined by Graf (1980). Images that did not fulfill these criteria were classified as “not a standard plane image” and the surgeons added relevant feedback. These non-standard images were included in the error analysis and the researchers reviewed the surgeons’ comments on images judged as non-standard. A second surgeon (AK) independently reviewed all cases, and any differences of opinion were resolved through discussion. Caregivers of infants with images suspicious of DDH were advised to seek early medical attention, while the outcomes of these referred cases were followed up.

### 2.6. Instrument Validity and Reliability

All screenings were conducted using the iViz air Ver.5 ultrasound system (Fujifilm, Tokyo, Japan). Before the study, pediatric orthopedic surgeons (TK, KA, KO, and KF) verified image quality. If more than three minutes were required for image capture, the PHN re-recorded the image. The same device was used in all sessions to ensure measurement consistency, and image quality was periodically reviewed by the research team during the study period.

### 2.7. Data Collection

PHNs’ demographic data were collected, including gender, ultrasound experience, DDH-screening training, number of HVs performed per month, and years of experience with newborn and infant HVs (Yoshioka-Maeda et al., 2024). The study was conducted as part of a local government program for newborn HVs, and informed consent was provided by both the administration and residents. Figure 2 illustrates the data flow.

**Figure 2.**
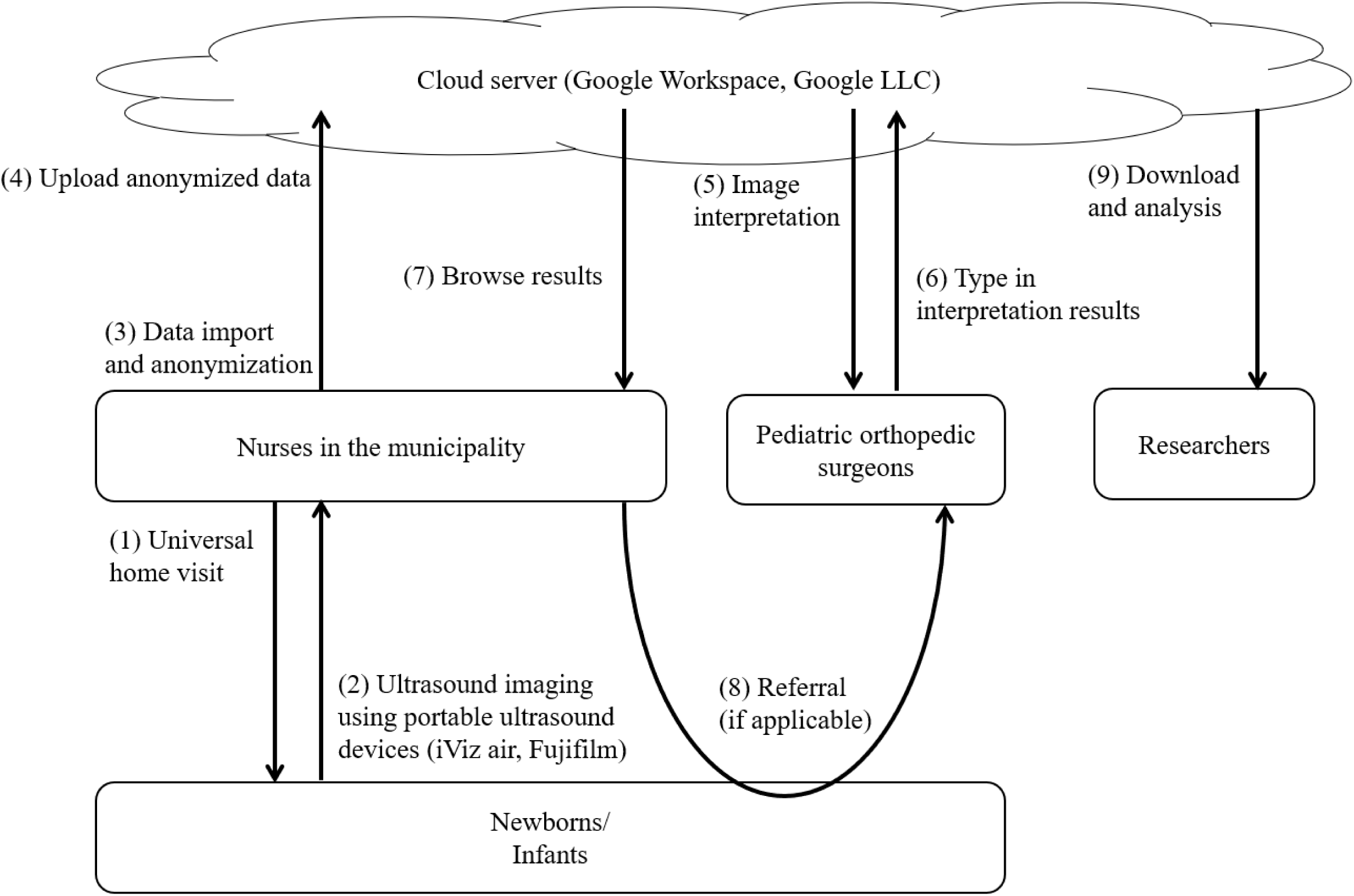
Data flow of the ultrasound hip screening during newborn and infant home visits

### 2.8 Demographic Characteristics of Newborns and Infants

The PHNs collected demographic information on the newborns and infants, including age and turn preference (positive, negative, or unknown). The presence or absence of the five DDH risk factors—female sex, breech birth, family history of DDH, asymmetric thigh, and groin creases, and limited abduction—was also recorded (Oka & Asagai, 2016).

### 2.9 Primary Outcomes

The primary outcome was the proportion of diagnostically interpretable images (%).

### 2.9 Secondary Outcomes

Secondary outcomes were imaging error patterns, immediate operational challenges, and follow-up results of infants with suspected DDH. Imaging-error patterns and immediate operational challenges were investigated using qualitative data. As noted above, pediatric orthopedic surgeons commented that unsuccessful images requiring re-imaging were “not standard images.” The additional comments were reviewed to classify common error patterns and examine how these related to the imaging process.

Additionally, the PHNs noted any difficulties encountered during image acquisition. Their field notes were used to identify operational challenges reflecting the feasibility of the intervention in real-world conditions. The follow-up results from referred cases were used to verify the appropriateness of the nurses’ initial image acquisition.

### 2.10. Analytic Strategy

After the opt-out period, the data were downloaded by the researchers, and Microsoft Excel (Microsoft Corporation, Redmond, WA, USA) was used to calculate the descriptive statistics of participants’ demographic and ultrasound-image characteristics.

Additionally, surgeons’ comments were thematically grouped to describe the distribution of imaging errors, while PHNs’ field notes were descriptively summarized to illustrate the operational challenges encountered during HVs. These findings were integrated to determine the screening’s overall technical and operational feasibility.

### 2.11. Ethical Considerations

This study was performed in accordance with the principles outlined in the Declaration of Helsinki and was approved by the Research Ethics Committee of our institution. The study’s procedure was explained to each caregiver using documents and a website. A six-week opt-out period followed each visit; only after this period were the data included in the study. No caregivers opted out of the study during this period.

## 3. Results

### 3.1. Sample Characteristics

All PHNs were female and none had previous ultrasound experience (Table 1). The PHNs visited 2.7 newborns/infants per month and had an average of 7.1 (standard deviation [SD]: 4.6) years of experience in performing HVs for newborns and infants.

**Table 1.**
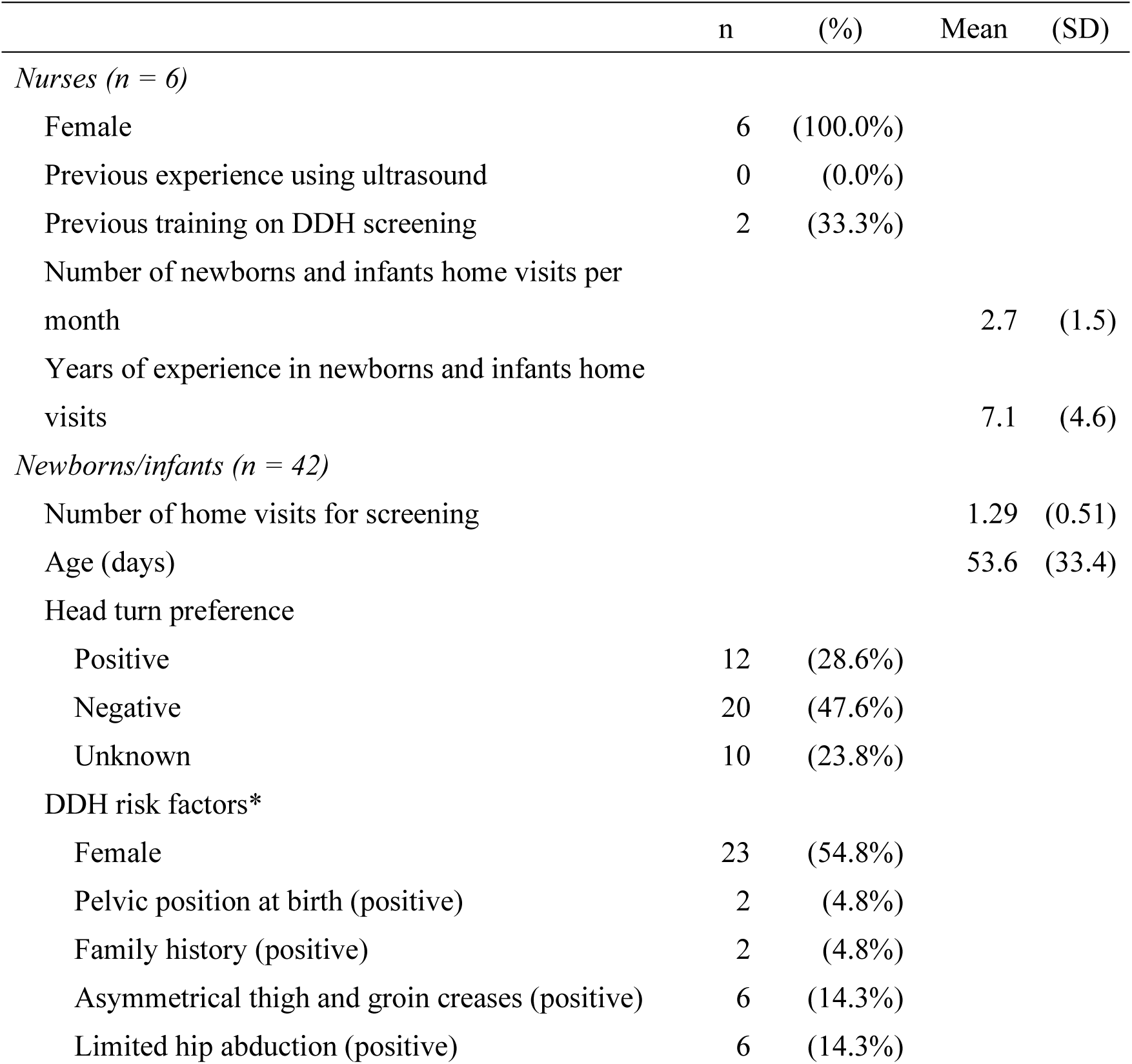
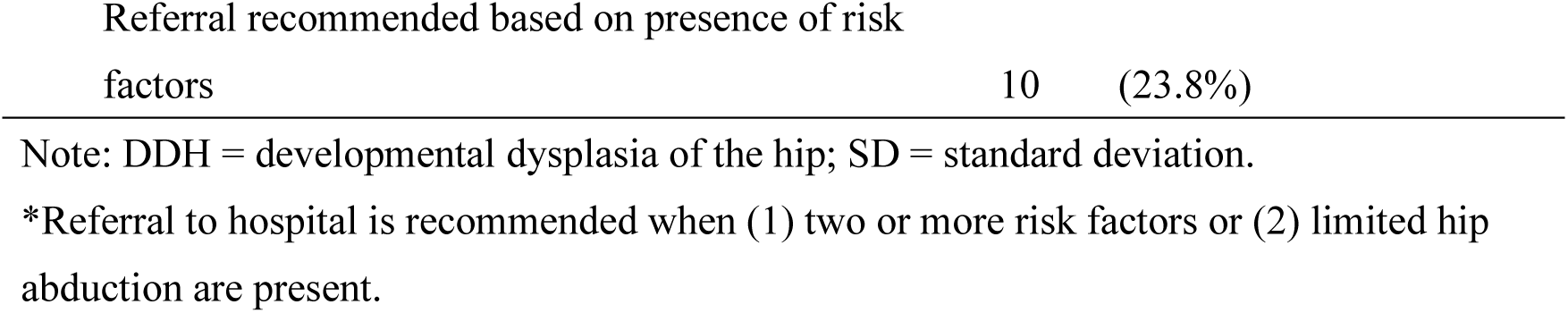
Nurse and newborn/infant characteristics.

The 42 children (19 boys and 23 girls) included in the study had a mean age (at the first HV) of 53.6 (SD: 33.4; range: 10–168) days; 1–3 HVs were performed (mean: 1.29). In terms of DDH risk factors, two children had pelvic birth positions, two had a family history of hip disease, six had asymmetrical thigh and groin creases, and six had limited abduction. Typically, referral to a hospital would be recommended when (1) two or more risk factors were present or (2) limited hip abduction was observed. Ten infants in the present study would have been referred to a hospital using this method.

### 3.2. Examination Process and Results

All infants eligible for newborn and infant HVs (n = 42) received ultrasound hip screening, indicating a high acceptability of implementation within the existing HV system. In the first HV, the PHNs successfully obtained diagnostically interpretable ultrasound images of 75 (89.3%), of a total of 84 hips. Nine hips’ (10.7%) images were judged by pediatric orthopedic surgeons as non-standard and required re-imaging. Since hip examinations were conducted during re-imaging after an initial imaging failure or when the surgeons ordered a follow-up examination, PHNs conducted 107 hip screenings. Thereafter, 12 (11.2%) hips were judged as non-standard. There were no significant differences between the diagnoses made by the two pediatric orthopedic surgeons.

Among these non-standard images, surgeons’ additional comments identified three main error patterns: incomplete visualization of the ilium (n = 2), incomplete visualization of the joint capsule (n = 1), curve of the bony roof (n = 2), and no detailed comment (n = 7). PHNs’ field notes revealed several operational challenges, including infant crying (n = 1). When examinations proved difficult, either multiple PHNs had to perform the screening (n = 1) or they had to inform the parents that the imaging would be performed at a later date (n = 1).

A summary of the screening results is presented in Table 2. The screening results showed that 33 infants (78.6%) had Type-I hips on both sides, four (9.5%) had Type-IIa hips on one or two sides, three (7.1%) had Type-IIc or-D hips, and two (4.8%) could not be evaluated because of a failure to capture standard images. Infants with Type-IIc or-D hips or those for whom standard images were not captured were referred to a hospital. Infants with imaging failures were older at the first screening (mean: 146.5 days), than compared with those for whom images were successfully captured.

**Table 2.** Screening results (n=42)

| Screening result | Screened |  | Potential<br>hospital<br>referral based<br>on risk factors | Actual<br>hospital<br>referral based<br>on screening | Visited<br>hospital | Treated | Age at screening<br>(days) |  | Number of home<br>visits for screening |  |
| --- | --- | --- | --- | --- | --- | --- | --- | --- | --- | --- |
|  | n | (%) | n | n | n | n | Mean | (Range) | Mean | (Range) |
| 1) Type I for both sides | 33 | (78.6%) | 6 | 0 | 0 | 0 | 51.8 | (10–117) | 1.2 | (1-2) |
| 2) Type IIa for both/one side | 4 | (9.5%) | 2 | 0 | 0 | 0 | 34.5 | (22–47) | 2.3 | (1-3) |
| 3) Type IIc or worse for both/one side | 3 | (7.1%) | 0 | 3 | 3 | 1 | 36.7 | (29–47) | 1.3 | (1-2) |
| 4) Imaging failure for both/one side | 2 | (4.8%) | 2 | 1 | 0 | 0 | 146.5 | (125–168) | 1.5 | (1-2) |

Note. Screening results indicate the final results after multiple home-visit examinations, if applicable. The infants with result 3) or 4) were to be
recommended to visit hospital.

Table 3 shows screening and follow-up histories. Of the 28 cases of Type-I hips, five underwent a second visit due to imaging failure at the first visit. Infants identified as having Type-IIa hips at the first visit received one or two additional follow-up visits at three-week intervals, along with health guidance, until regression to Type-I hip was confirmed. One infant identified as having Type-IIa hips at the first visit was suspected to have Type-D hips at the second visit; however, assessment at the hospital determined that the infant was normal. The infant identified as having a Type-IIc hips at the first screening visit was diagnosed with Type IIc by a pediatric orthopedic surgeon at a hospital; two months later, both sides were normal. Another infant, identified as having Type-D hips, was diagnosed with Type-D hips by a pediatric orthopedic surgeon and is currently undergoing orthotic treatment. One infant with imaging failure was not referred to a hospital but was assessed as normal by a pediatrician at the routine nine-month check. A hospital assessment was recommended for the remaining infant with imaging failure. One infant had not visited the hospital as of the writing of this study.

**Table 3.** Screening and follow-up histories (n=42)

| Category | n | First visit | Second visit | Third visit | Medical examination results |
| --- | --- | --- | --- | --- | --- |
| 1) Type I for both sides | 28 | Type I for both sides | - | - | - |
|  | 5 | Type I & imaging failure | Type I for both sides | - | - |
| 2) Type IIa for both/one side | 1 | Type IIa for both sides | Type I for both sides |  | - |
|  | 2 | Type IIa & Type I | Type I for both sides |  | - |
|  | 1 | Type IIa & Type I | Type IIa for both sides & Type I for one side | Type I for both sides | - |
| 3) Type IIc or worse for both/one side | 1 | Type IIa & Type I | Type D & Type I | - | Assessed as normal on both sides by a pediatric orthopedic surgeon. |
|  | 1 | Type IIc & Type IIa | - |  | Assessed as Type IIa & Type IIc by a pediatric orthopedic surgeon; assessed as normal on both sides two months late. |
|  | 1 | Type D & Type IIa | - |  | Assessed as Type D & Type IIa by a pediatric orthopedic surgeon; orthotic treatment ongoing. |
| 4) Imaging failure for both/one side | 1 | Imaging failure for both sides | Imaging failure for both sides | - | None. This infant (caregiver) was recommended to visit a hospital but did not. |
|  | 1 | Imaging failure for both sides | - | - | Assessed as normal on both sides by a pediatrician at the routine nine-month check |

Finally, three of the four infants recommended for a visit were examined by a pediatric orthopedic surgeon (TK) at the hospital, while one was treated with an orthosis. None of the infants referred to the hospital due to imaging findings of Type-IIc hips or worse, including the treated infant, would have been referred using the risk-factor method.

## 6. Discussion

This study investigated whether trained PHNs could obtain diagnostically interpretable ultrasound hip images during HVs, demonstrating the feasibility and added value of integrating nurse-led DDH screening with existing maternal and child health programs. This approach may enhance early DDH detection in communities where access to pediatric orthopedic specialists is limited.

The infants underwent ultrasound screening at a mean age of eight weeks, with follow-up screening conducted three weeks later. Although some European countries conduct screening shortly after birth (Kilsdonk et al., 2021), no global consensus exists regarding such screenings’ optimal timing, partly because physiological ligamentous laxity is influenced by maternal estrogen levels in infants younger than six weeks (American Institute of Ultrasound in Medicine, 2024). Ongoing debates on universal versus selective screening to avoid overtreatment (Agostiniani et al., 2020; Kuitunen et al., 2022) and operator-dependent image quality (Graf et al., 2013), highlight the need for structured user training (Duarte et al., 2022). Nevertheless, PHNs in this study successfully produced interpretable images in most cases, thus offering supporting evidence of the practicality of ultrasound hip screening during HVs.

Among seven infants classified as having Type-IIa hips or worse, five would have been overlooked by risk-factor-based screening, including one requiring orthotic treatment. This underscores the limited sensitivity of physical examinations and risk-based screening, which often fail to detect bilateral or subtle DDH cases (Harper et al., 2020; Smart et al., 2024). Therefore, incorporating ultrasound screenings into HV programs may prevent delayed DDH diagnoses. In Japan, early conservative management—typically, Pavlik harness therapy is performed in outpatient settings before patients are six months old—costs approximately 20,500 yen, whereas open surgical treatment for late-diagnosed DDH costs approximately 232,400 yen (Ministry of Health, Labour and Welfare, 2024)—nearly 11 times higher. Early detection DDH improves gait, quality of life (Shorter et al., 2013), and reduces future osteoarthritis risk (Sato et al., 2024). Therefore, nurse-led ultrasound hip screening may contribute to long-term cost savings.

In Japan, maternal and child health services are administered by local governments and function separately from medical facilities. Because preventive ultrasound screening is not reimbursed by national insurance, municipalities depend on pediatric orthopedic surgeons’ voluntary support for interpretation and follow-up.

Additionally, workforce shortages limit the availability of community-based screening (Yoshioka-Maeda et al., 2023). Against this backdrop, nurse-led ultrasound can complement physician-led screening and expand access; however, a sustainable budgeting and reimbursement framework is essential, especially in rural areas.

Although children who required repeat visits for Type-IIa findings generally showed improvement without medical treatment, PHNs also provided guidance on proper diapering, clothing, and baby-wearing—practices that support healthy hip development and DDH prevention (Ishida, 1977; Yamamuro & Ishida, 1984). This dual function of PHNs as both screeners and educators highlights the importance of combining ultrasound assessment with tailored preventive care during HVs.

Two cases (4.8%) were classified as inadequate because of non-standard images.

Such failures may stem from technical factors or true hip instability that prevents the femoral head from remaining in the acetabulum. As Graf (2006) notes, suboptimal images may make normal hips appear abnormal; however, proper imaging cannot make abnormal hips appear normal—thus, false positives are more likely than false negatives.

In this study, one inadequate case was later judged as normal, while the other required referral, suggesting that repeated imaging failures should prompt clinical suspicion of instability rather than be dismissed as an operator error. With pediatric orthopedic surgeons interpreting all images, nurse-led ultrasound screening appears to be a safe and feasible strategy for early DDH detection in home settings.

### 4.1. Strengths and Limitations

PHNs conducted ultrasound DDH screening during newborn and infant HVs. Performing ultrasonography in diverse home environments is challenging; however, caregivers cooperated to keep the infants in a lateral position, similar to hospital conditions. PHN-led ultrasound screening combined with preventive health guidance could improve physical-assessment accuracy during HVs, contributing to early DDH detection and prevention. Traditional physical examinations often overlook DDH (Harper et al., 2020), which underscores the need for accuracy when conducting assessments in community settings (International Council of Nurses, 2021).

This study had certain limitations. First, it was conducted in a single municipality, which limits its findings’ generalizability. Second, the PHNs conducted additional HVs for Type-IIa cases based on the advice of pediatric orthopedic surgeons.

In this municipality, PHNs routinely perform multiple HVs; however, this may be difficult to implement elsewhere due to limited resources. Third, it is necessary to improve PHNs’ technical ability to enhance screening accuracy.

### 4.2. Recommendations for Further Research

Future studies should include larger and more diverse populations from different regions. Cost–benefit analyses are required, while early DDH detection’s long-term impact should be evaluated. Additionally, to increase imaging success rates, it is necessary to refine training programs for PHNs with limited ultrasound experience.

Implementing artificial intelligence to assist in the assessment of suspicious imaging findings may also support early referral.

### 4.3. Implications for Policy and Practice

Early DDH detection is a global priority (Agostiniani et al., 2020). Newborn and infant HVs provide an opportunity for concurrent screening and caregiver education at home, thereby fostering healthy hip development. As nurse-led HVs are conducted worldwide, they can contribute significantly to early DDH detection. From a cost-reduction perspective, improving community-based screening accuracy is crucial.

National and local governments should support nurse-led ultrasound hip screening during HVs for newborns and infants as a novel and precise public-health nursing model. Establishing remote diagnostic systems that facilitate collaboration with pediatric orthopedic surgeons and subsidizing the acquisition of portable ultrasound devices could enhance early DDH detection in community settings.

### 4.4. Conclusion

Nurse-led universal ultrasound hip screening during newborn and infant HVs can enable early DDH detection. Of the 42 infants, approximately 80% had normal hips, approximately 10% required detailed examinations at medical facilities because of abnormal findings, and approximately 10% experienced improved hip condition following the implementation of abduction exercises to prevent hip dislocation. PHNs successfully collaborated with pediatric orthopedic surgeons and caregivers to perform universal ultrasound screening and provide preventive health guidance; this dual role could revolutionize early DDH detection in community settings. By combining ultrasound hip screening with preventive health guidance, PHNs can enhance the precision of public-health nursing and the value of HVs for newborns and infants, potentially contributing to the prevention of gait disorders, improved quality of life, and reduced healthcare costs.

## Data Availability

Research data are not publicly shared because of privacy restrictions.

